# Baseline severity of positive and negative symptoms predicts relapse in opposing directions among placebo-treated schizophrenia patients: analysis of five placebo-controlled, antipsychotic withdrawal, relapse-prevention trials

**DOI:** 10.64898/2026.09.28.26363883

**Authors:** Jonathan Rabinowitz, Michael Davidson, Christoph U. Correll, Helene Speyer, Mattan S. Ben-Shachar, Adam Noy, Anamaria Ciubară, Remy Luthringer

## Abstract

**Background and Hypothesis:** Identifying which patients with schizophrenia will remain stable and which will relapse is relevant to both trial design and clinical practice. We hypothesized that higher negative and lower positive symptoms at baseline would predict longer time to relapse.

**Study Design:** This analysis pooled individual participant data on 557 patients with schizophrenia randomized to placebo from five placebo-controlled antipsychotic withdrawal relapse-prevention studies, each with a double-blind phase of at least 6 months. Relapse was defined as a ≥12-point increase in Positive and Negative Syndrome Scale (PANSS) total score from double-blind baseline at any time during the double-blind phase. Cox regression analyses modeled time to relapse with the three PANSS subscales, age and sex.

**Study Results:** PANSS Negative and Positive symptom subscales predicted relapse in opposite directions and at almost equal magnitude in the pooled model. Higher negative symptoms predicted lower relapse risk (hazard ratio (HR)=0.94 per point, 95% confidence interval (CI)=0.90-0.99) and higher positive symptoms predicted higher relapse risk (HR=1.06, 95% CI=1.00-1.13). PANSS General Psychopathology did not predict relapse (HR=0.98, 95% CI=0.95-1.02). Based on the pooled estimates, a 4-point higher baseline PANSS Positive score corresponded to a 27% higher relapse risk, while a 4-point higher baseline PANSS Negative score corresponded to a 21% lower risk.

**Conclusions:** These results may help predict relapse risk in clinical care and in clinical trial samples but require prospective validation.

## Introduction

Several meta-analyses have reported that, on average, maintenance antipsychotic treatment reduces relapse risk in schizophrenia, with roughly a threefold difference in one-year relapse rates between drug and placebo groups in randomized withdrawal designs.^1^ However, this average effect conceals substantial variation: some patients randomized to placebo remain clinically stable for a year or longer. Identifying these patients in advance matters for patients and clinicians who might want to pursue targeted or intermittent antipsychotic treatment strategies^2,3^ to reduce the short-and long-term adverse effects of dopamine receptor blocking drugs^4–6^ while minimizing the risk of potentially detrimental effects of relapse.^7^

This issue is also important for trial design. A relapse-prevention trial can demonstrate a treatment effect only if enough relapse events occur; therefore, a sample drawn from a population at low relapse risk may leave the trial underpowered. The opposite problem arises in trials targeting negative symptoms or cognition: relapse is not the outcome of interest but a competing event and a major source of attrition. In such trials, enrolling a high-risk sample can undermine the study aims.^8^

In a reanalysis of individual participant data from randomized discontinuation studies of oral and long-acting injectable antipsychotics, lower baseline symptom severity and longer prior stability predicted a lower likelihood of relapse.^9^ In phase 1 of the Clinical Antipsychotic Trials of Intervention Effectiveness (CATIE), 25.5% of 1,139 chronically ill participants met a 12-point Positive and Negative Syndrome Scale (PANSS) worsening criterion for relapse over 18 months, and those in the medium and high baseline negative symptom tertiles were at lower risk of worsening than those in the lowest tertile (hazard ratio (HR)=0.73 and HR=0.69, respectively).^10^ Converging evidence comes from clinical studies of roluperidone, in which patients with moderate-to-severe negative symptoms treated with roluperidone alone (a 5-HT2A, sigma-2 and alpha-1A antagonist without dopamine receptor blockade) showed low rates of symptomatic worsening over 12 months.^11^

This evidence is limited because participants were taking antipsychotics. In a medicated cohort, an association between high baseline negative symptoms and low risk of worsening can be read in two ways. This relationship may reflect the natural course of an illness dominated by deficit features, or it may be a product of treatment: antipsychotics can cause secondary negative symptoms,^4,6^ and patients with prominent negative symptoms may differ in adherence, in antipsychotic dose, or in how their positive symptoms respond to dopamine blockade. Separating the two scenarios requires observing stabilized patients who are not taking an active psychotropic compound. Opportunities to do so are scarce. Almost all placebo-controlled trials in schizophrenia are acute-phase studies lasting 4-12 weeks,^12^ too brief for relapse to serve as the outcome, while naturalistic discontinuation cohorts lack blinding and standardized ratings and are subject to confounding by indication, since patients whose clinicians discontinue their medication do not constitute a random subset. The placebo arms of double-blind, placebo-controlled, antipsychotic withdrawal, relapse-prevention trials are the one setting that avoids both problems: participants are stabilized on an antipsychotic, withdrawn to placebo under blinded conditions, and assessed with standardized symptom ratings at scheduled visits for months rather than weeks.

The trials analyzed here, conducted between 2004 and 2014, constitute a unique body of evidence on the untreated course of stabilized schizophrenia; we were unable to identify other such trials with available participant-level data. We pooled individual participant data from the placebo arms of these five similarly designed trials, each with a double-blind phase of at least 6 months, to examine whether baseline psychopathology at entry into the double-blind phase predicts time to relapse. The hypothesis and analysis plan were specified in advance, in data requests submitted to the two data-sharing platforms before any data were released.^13,14^ We examined the independent contributions of the positive, negative and general psychopathology domains and hypothesized that participants with predominantly negative symptoms and correspondingly low positive symptoms at baseline would remain relapse-free longer than participants without this profile.

## Methods

### Data sources and study selection

Individual participant data were obtained through two independent data-sharing platforms, Vivli and the YODA Project, under executed data use agreements. Both data request applications set out the objective, hypothesis, and outcome definitions and the analysis plan before any data were released, and both were publicly posted^13,14^; the analyses reported here follow those plans. Studies were eligible if they (i) used a randomized, double-blind, placebo-controlled relapse-prevention or randomized-withdrawal design in adults with schizophrenia; (ii) had a double-blind phase of at least 6 months; (iii) administered the PANSS^15^ at entry to the double-blind phase and at scheduled visits thereafter; and (iv) made participant-level data available through either platform. Five studies met all criteria (Table 1): four Janssen relapse-prevention trials accessed through the YODA Project,^16–19^ and one Eli Lilly trial accessed through Vivli.^20^ YODA Project data were made available on the Vivli platform, where all analyses were conducted.

**Table 1.** Studies contributing placebo-randomized participants and baseline characteristics by study.

| Study, registration | Placebo n | Women n (%) | Age M (SD) | PANSS-P M (SD) | PANSS-N M (SD) | PANSS-G M (SD) | PANSS total M (SD) |
| --- | --- | --- | --- | --- | --- | --- | --- |
| 1. NCT00111189 | 180 | 84 (46.7) | 41.0 (10.9) | 11.0 (3.1) | 14.9 (4.6) | 26.7 (6.2) | 52.7 (11.9) |
| 2. NCT00086320 | 74 | 28 (37.8) | 38.1 (11.1) | 11.9 (3.8) | 14.9 (3.8) | 28.4 (7.5) | 55.2 (13.1) |
| 3. NCT01662310 | 67 | 38 (56.7) | 32.0 (12.3) | 11.7 (4.4) | 15.6 (4.5) | 27.2 (7.4) | 54.4 (13.9) |
| 4. NCT01529515 | 136 | 33 (24.3) | 36.4 (11.4) | 11.3 (2.9) | 16.1 (3.9) | 26.5 (5.0) | 53.9 (9.2) |
| 5. - | 100 | 47 (47.0) | 35.1 (11.2) | 9.0 (2.4) | 11.9 (4.0) | 22.2 (5.4) | 43.0 (10.7) |
| Total | 557 | 230 (41.3) | 37.3 (11.6) | 10.9 (3.4) | 14.7 (4.4) | 26.1 (6.4) | 51.8 (12.2) |

Note. PANSS-P, PANSS-N and PANSS-G = Positive, Negative and General Psychopathology subscales of the Positive and Negative Syndrome Scale, measured at entry into the double-blind phase. Placebo n is the number of placebo-randomized participants contributing to the survival analysis. Race was coded inconsistently across datasets and is not reported; 44 participants (7.9%) were recorded as Hispanic or Latino. Open-label period and stability criteria required for randomization, by study: (1) 9-week transition plus 24-week maintenance (33 weeks); PANSS total $\leq 75$ and items P1, P2, P3, P6, P7, G8 and G14 each $\leq 4$ . (2) 8-week run-in plus 6-week stabilization (14 weeks); PANSS total $\leq 70$ , study 1 items except G14 each $\leq 4$ , and CGI-S $\leq 4$ . (3) 8-week run-in plus 6-week stabilization (14 weeks). (4) 17-week transition (PANSS total $< 70$ required to continue) plus 12-week maintenance (29 weeks); PANSS total $< 70$ and study 1 items each $\leq 4$ at weeks 21, 25 and 29. (5) 6-week conversion plus 8-week stabilization on olanzapine (14 weeks). The double-blind phase was of variable duration (until relapse, withdrawal or study termination) in studies 1–4 and planned for up to 52 weeks in study 5. Protocol ID and sponsor, by study: (1) R092670PSY3001 (CR004198), Janssen; (2) R076477-SCH-301 (CR004384), Janssen; (3) R076477-SCH-3041 (CR100427), Janssen; (4) R092670PSY3012 (CR100717), Janssen; (5) F1D-MC-HGGI, Eli Lilly. Source: Studies 1-5 YODA, 6 VIVILI

### Participants

The analysis sample comprised the 557 participants randomized to placebo across the five studies. All participants had been stabilized during an open-label run-in of protocol-specified duration that ranged from 14 to 33 weeks and had met each parent protocol’s stability criteria before double-blind randomization; run-in duration and stability definitions varied across studies and are detailed in the Table 1 notes. Using the study numbers in Table 1, the open-label period was 14 weeks in studies 2, 3 and 5, 29 weeks in study 4 and 33 weeks in study 1.

### Measures and outcome

In each of the five trials, patients were first treated for an acute exacerbation until they met predefined stability criteria. Individuals then entered a double-blind maintenance phase where they either received an antipsychotic or, after antipsychotic withdrawal, placebo. Baseline was defined as the last PANSS assessment on or before randomization into the double-blind maintenance phase. The outcome was time from double-blind randomization to relapse, defined as an increase of 12 points or more from double-blind baseline on the PANSS total score. This criterion was chosen in preference to the sponsor-defined relapse endpoints of the individual protocols, which combined symptom thresholds with hospitalization, clinical global impression change, suicidality and aggression in study-specific ways and are therefore not comparable across trials. The 12-point threshold is scale-based, uniformly computable from the raw item data, and has been validated against clinician-rated relapse and hospitalization.^21^

### Statistical analysis

Time to relapse was modeled with Cox proportional-hazards regression analyses. Baseline PANSS Positive, Negative and General Psychopathology subscale scores were entered simultaneously as continuous predictors together with age (in years) and sex. Entering the three subscales together estimates each domain conditional on the other two. The proportional-hazards assumption was assessed with the global test based on scaled Schoenfeld residuals.

The hypothesis was that both associations hold: higher PANSS positive symptoms significantly raise relapse risk, and higher PANSS negative symptoms significantly lower it. This hypothesis was tested with an intersection-union test, which requires both associations to be significant in the prespecified directions. The *P* value is the larger of the two one-sided component *P* values; no extra multiplicity adjustment is needed because this test is already conservative. The directions were specified before the data were released.^13,14^ We also tested whether the positive and negative symptom effects were equal in size but opposite in direction. If so, the two terms could be summarized by one score: Positive minus Negative symptoms, with General Psychopathology, age and sex kept in the model.

The one-stage model assumes a common baseline hazard across trials that differ appreciably in baseline severity and design. The analysis was therefore repeated in two stages: the Cox model was fitted separately within each of the five studies, and the resulting study-specific log hazard ratios were combined by random-effects meta-analysis. The two-stage estimate uses only within-study variation, so it avoids the aggregation bias that can arise when studies with different mean symptom levels and different event rates are pooled into one model. Both estimates are reported. Hazard ratios are reported with 95% confidence intervals. Because the direction of each association was prespecified before data release, *P* values for the directional hypotheses (the baseline PANSS Positive and Negative subscales and the Positive-minus-Negative difference score) are one-sided; all other *P* values, including those for covariates and for the proportional-hazards and coefficient-equality tests, are two-sided. Alpha was set at .05. Because a per-point hazard ratio is difficult to interpret at the scale on which eligibility criteria are actually written, we also report the hazard ratio implied by the fitted model for between-patient differences of one to four points on a subscale, computed as the per-point hazard ratio raised to the power of the score difference. For comparability with studies reporting effects per standard deviation or across score tertiles, hazard ratios were also rescaled to the pooled baseline standard deviation of each subscale. To display the fitted association on the survival scale, model-implied survival curves were generated by the pick-a-point method at baseline PANSS Negative scores of 10, 15 and 20, spanning approximately one standard deviation on either side of the pooled mean of 14.7, with age and the Positive and General Psychopathology subscales held at their sample means, and sex held at its modal value. Analyses were conducted in R version 4.4.0 using the survival package.

## Results

### Predictors of time to relapse

For a description of the studies and the analyzed sample see Table 1. Overall, 32.3% of patients relapsed during double-blind follow-up; among those who relapsed, the mean time to relapse was 9.75±8.52 weeks (Supplemental Figure 1). In the Cox model (Table 2; Supplemental Figure 2), the PANSS Positive and Negative symptom domains were associated with relapse in opposing directions and at nearly equal magnitude. Each additional point on the baseline PANSS Negative subscale was associated with a 6% lower hazard of relapse (b = -0.058, HR = 0.944, 95% CI: 0.902-0.989, one-sided *P* = .007), and each additional point on the baseline PANSS Positive subscale was associated with a 6% higher hazard (b = 0.060, HR = 1.061, 95% CI: 0.997-1.129, one-sided *P* = .030). The two coefficients were close to mirror images of one another. PANSS General Psychopathology scores were unrelated to relapse (HR = 0.984, 95% CI 0.946-1.023, *P* = .42). Relapse risk was therefore associated with the balance between positive and negative symptoms. Among the examined covariates, older age predicted lower relapse risk (HR = 0.986 per year, 95% CI 0.974-0.999, *P* = .036), equivalent to a 13% lower hazard per decade. The proportional-hazards assumption was not violated (global test χ^2^(5) = 3.57, *P* = .61).

**Table 2.** Cox proportional-hazards model of time to relapse among placebo-randomized participants (N = 557).

| Baseline predictor | b (SE) | HR | 95% CI | P | HR per SD |
| --- | --- | --- | --- | --- | --- |
| PANSS Positive (per 1 point) | 0.060 (0.032) | 1.061 | 0.997-1.129 | .030 | 1.22 |
| PANSS Negative (per 1 point) | -0.058 (0.023) | 0.944 | 0.902-0.989 | .007 | 0.78 |
| PANSS General Psychopathology (per 1 point) | -0.016 (0.020) | 0.984 | 0.946-1.023 | .42 | 0.90 |
| Age (per year) | -0.014 (0.006) | 0.986 | 0.974-0.999 | .036 | - |
| Sex (male vs. female) | -0.254 (0.152) | 0.776 | 0.576-1.045 | .095 | - |

Note. b = regression coefficient; SE = standard error; HR = hazard ratio; CI = confidence interval; SD = standard deviation. P values for the PANSS Positive and Negative subscales are one-sided, reflecting the prespecified directional hypotheses; all other P values are two-sided. All predictors were entered simultaneously; the model was fitted to the pooled sample with a common baseline hazard across studies. HR per SD rescales the estimate to the pooled baseline standard deviation of the subscale (3.4 points for Positive, 4.4 for Negative, 6.4 for General Psychopathology). The global test of the proportional-hazards assumption was not significant, $\chi^2(5) = 3.57$ , $P = .61$ .

### Testing the opposing-directions hypothesis

The prespecified joint hypothesis, that both associations hold, was supported by the intersection-union test (*P* = .03; one-sided component *P* values: .03 for the PANSS Positive subscale and .007 for the PANSS Negative subscale). The Positive and Negative coefficients were nearly equal in size and opposite in sign (0.060 vs. -0.058; test of equal magnitude, z = 0.05, *P* = .96). We therefore combined them into a single difference score (PANSS Positive minus PANSS Negative), which gave a hazard ratio of 1.06 per point (95% CI: 1.01-1.10, one-sided *P* = .004).

### Two-stage random-effects meta-analysis

As a check on the pooled result, we repeated the analysis separately within each of the five trials and then combined the five trial-level results. In this analysis, the estimate for baseline PANSS Negative symptoms was weaker: each 1-point higher Negative score was associated with an estimated 3% lower relapse hazard (HR = 0.97 per point, 95% CI: 0.91-1.02; b = -0.030, z = - 1.19, one-sided *P* = .116), as shown in Figure 3. For baseline PANSS Positive symptoms, the two-stage estimate was stronger than in the pooled model (HR = 1.13 per point, 95% CI: 1.06-1.21; b = 0.12, z = 3.52, one-sided *P* = .0002). The Negative estimate thus pointed in the same direction as the pooled model, but it was smaller and its confidence interval included 1 (no effect).

The individual trials were broadly consistent with a small and uncertain Negative-symptom effect. Three trials estimated lower relapse risk with higher Negative scores, with hazard ratios of 0.92 (95% CI: 0.82-1.03), 0.93 (0.85-1.03) and 0.94 (0.84-1.05). The other two trials did not show this pattern, with hazard ratios of 1.01 (0.82-1.25) and 1.04 (0.95-1.14). However, none of the individual trials gave a precise result, and their confidence intervals overlapped substantially. Thus, the trial-level results are compatible with a modest protective association, but no single trial establishes it on its own.

### Magnitude of the associations

To make the size of the effects easier to compare, we also expressed them per standard deviation of each subscale. On this scale, a 1-standard-deviation higher Negative score was associated with a lower relapse hazard (HR = 0.78, 95% CI: 0.63-0.95), while a 1-standard-deviation higher Positive score was associated with a higher relapse hazard (HR = 1.22, 95% CI: 0.99-1.50). Thus, a 1-standard-deviation difference in negative symptoms corresponded to about a 22% lower relapse hazard, and a 1-standard-deviation difference in positive symptoms to about a 22% higher relapse hazard. The two effects were therefore similar in size but pointed in opposite directions. In the more conservative two-stage analysis, the Negative estimate was weaker (HR = 0.88 per standard deviation, 95% CI: 0.67-1.10) and its confidence interval included 1; the Positive estimate was stronger (HR = 1.51 per standard deviation, 95% CI: 1.20-1.91).

These per-point effects accumulate across larger score differences. For example, as shown in Figure 1, a 4-point higher Negative score was associated with a lower relapse hazard (HR = 0.79, 95% CI: 0.66-0.96), while a 4-point higher Positive score was associated with a higher relapse hazard (HR = 1.27, 95% CI: 0.99-1.63), with the other variables held constant. A patient who was 4 points higher on Positive symptoms and 4 points lower on Negative symptoms than another patient would have an estimated relapse hazard about 1.6 times as high. Figure 2 shows the same association on the survival scale: model-implied time-to-relapse curves at baseline PANSS Negative scores of 10, 15 and 20 separate progressively over the maintenance phase, with higher Negative scores corresponding to a lower cumulative probability of relapse.

**Figure 1.**
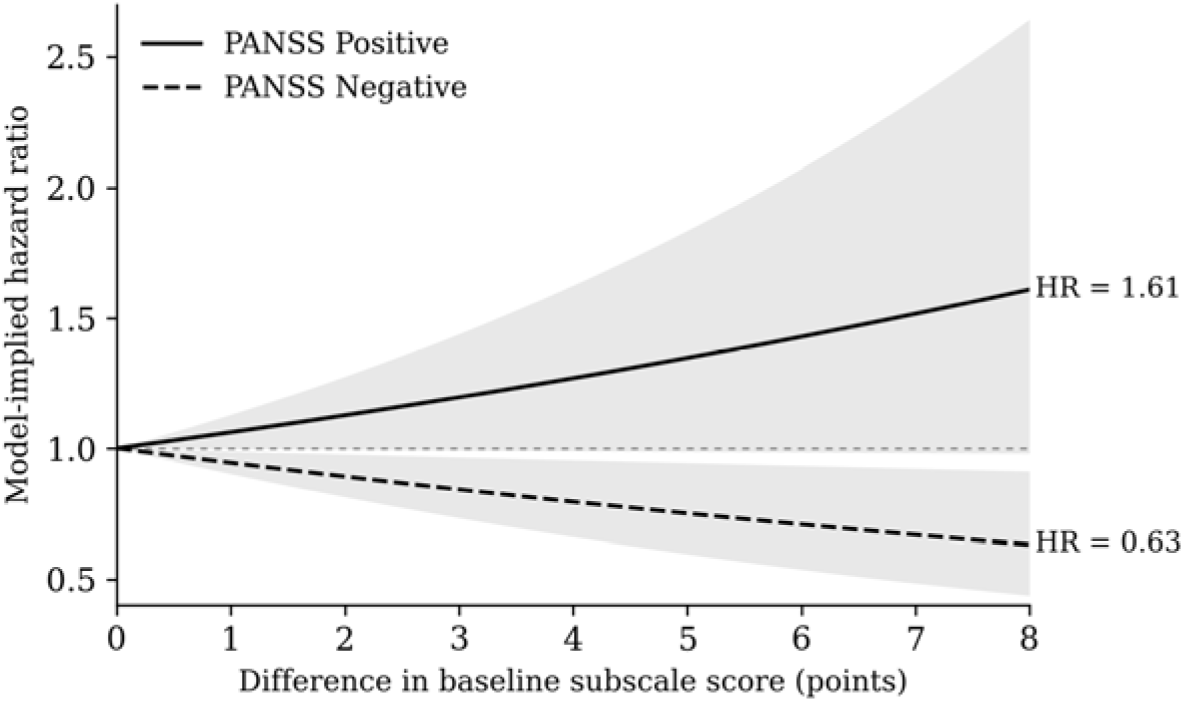
Hazard ratios implied by the fitted model for between-patient differences of 1 to 4 points on a baseline PANSS subscale, holding the remaining covariates constant. At a 4-point difference the implied hazard ratio is 0.79 (95% CI 0.66-0.96) for the Negative subscale and 1.27 (95% CI 0.99-1.63) for the Positive subscale. Shaded bands are 95% confidence intervals.

**Figure 2.**
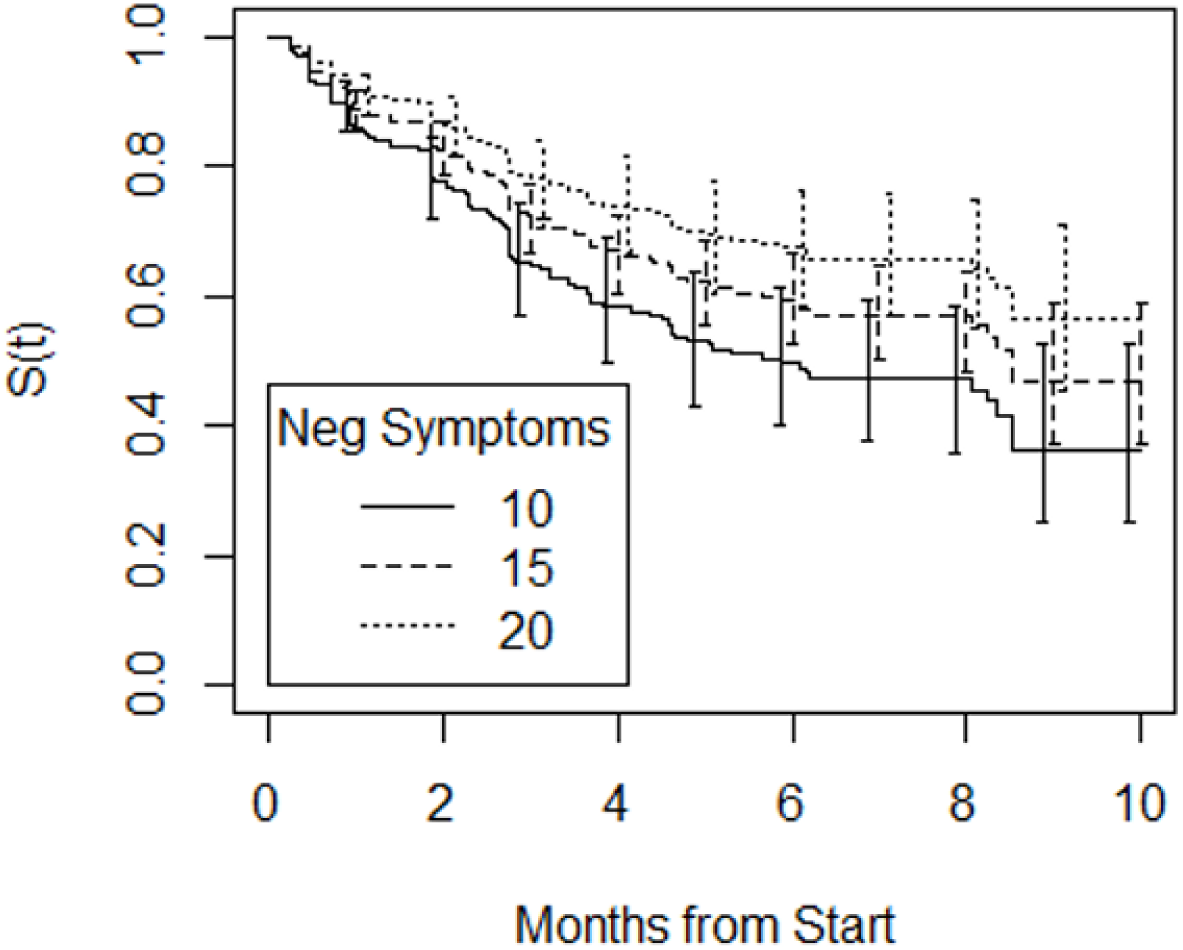
Model-implied survival functions for time to relapse at baseline PANSS Negative subscale scores of 10, 15 and 20, with age and the Positive and General Psychopathology subscales held at their sample means and sex held at its modal value. Dashed vertical lines are pointwise 95% confidence intervals. Higher baseline negative symptom scores correspond to higher relapse-free survival throughout follow-up.

**Figure 3.**
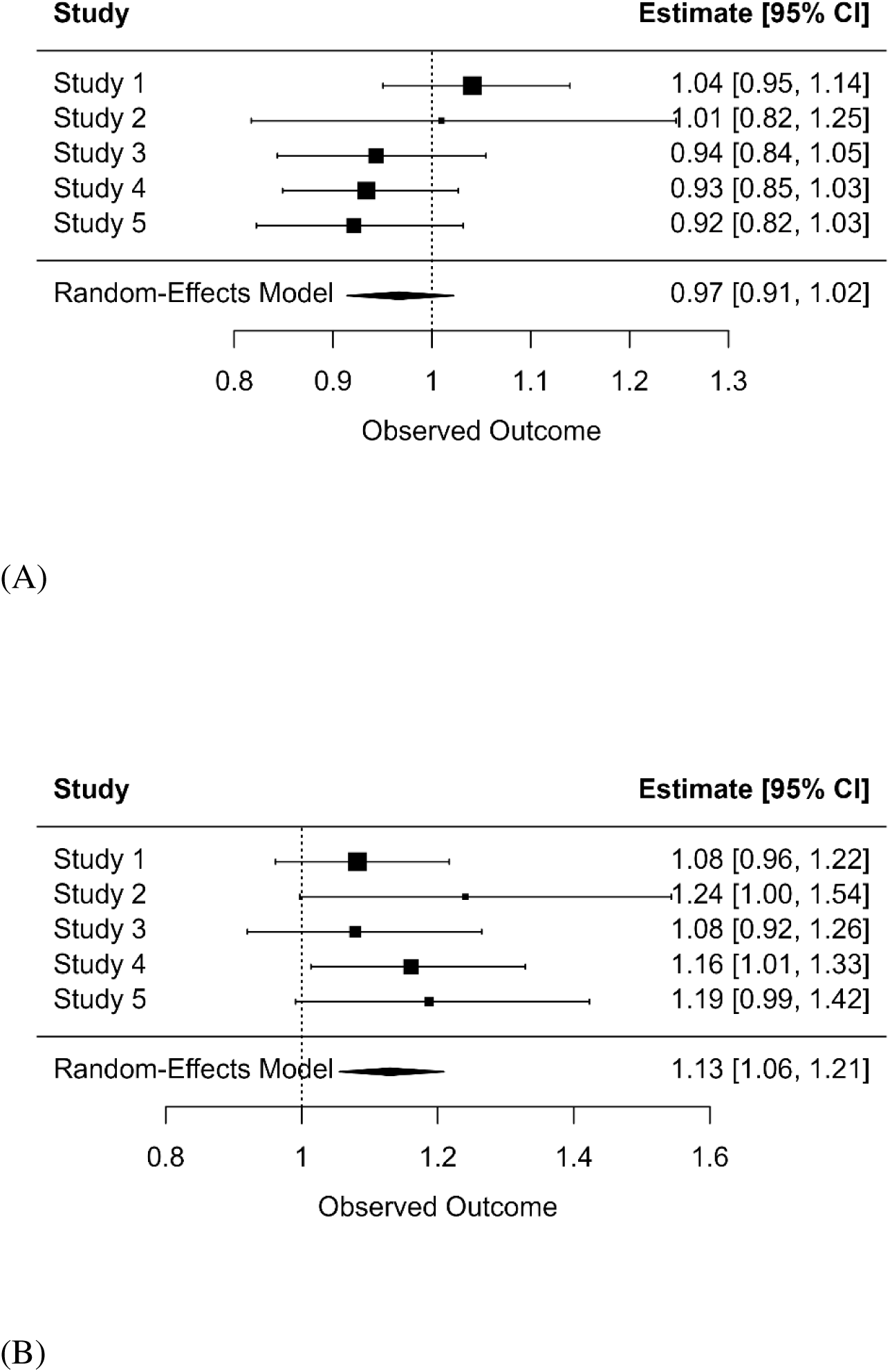
Forest plots of study-specific hazard ratios for the baseline PANSS Negative (A) and Positive (B) subscales, per 1-point higher score, with the pooled random-effects estimate. Squares denote study-specific point estimates and horizontal lines their 95% confidence intervals; the diamond denotes the pooled estimate and its confidence interval. Marker size is proportional to study weight.

## Discussion

This analysis included 557 multi-episode patients with schizophrenia who were stabilized on antipsychotic treatment after an acute exacerbation and then randomized to placebo under double-blind conditions. In this sample, baseline positive and negative symptoms predicted time to relapse in opposite directions and, in the pooled model, with nearly identical effect sizes. Each 1-point higher baseline positive score was associated with a 6% higher relapse hazard, whereas each 1-point higher baseline negative score was associated with a 6% lower relapse hazard. General psychopathology provided no additional prognostic information after accounting for the other two domains. Model-implied survival curves separated progressively over the maintenance phase (Figure 2).

These findings align with a post-hoc analysis of the CATIE phase-1 data showing that participants with medium or high baseline negative symptoms had a lower risk of 12-point PANSS worsening than those with low negative symptoms (HR = 0.73 and 0.69).^10^ In the present placebo-treated sample, the Negative symptom estimate was similar in direction and magnitude when expressed per standard deviation (HR = 0.78). Together, these results suggest that the association between higher negative symptoms and lower relapse risk is not merely an artifact of medication exposure, because it was also observed among patients receiving placebo.

The CATIE comparison and the present placebo analysis point in the same direction, but the evidence is not decisive. The strongest conclusion is that the association is reproducible enough to merit attention and prospective validation, while its exact size remains uncertain. Negative symptoms, particularly enduring (primary) negative symptoms, are associated with a less fluctuating course and with lower reactivity of the dopaminergic systems that mediate psychotic exacerbation.^22,23^ On this reading, the association between higher negative symptoms and lower relapse risk is prognostic in a clinically meaningful sense.

These findings differ from parts of the first-episode literature, where baseline negative symptoms have been associated with higher relapse rates regardless of treatment strategy.^24–26^ The two sets of findings are not necessarily contradictory. In first-episode illness, when patients are typically young, negative symptoms may indicate an emerging trajectory of greater severity; in stabilized, chronically ill patients, they may instead identify a course marked by fewer acute fluctuations, resembling what was historically described as “residual schizophrenia.” Thus, the prognostic meaning of negative symptoms may depend on illness stage, a possibility that requires further investigation.

Relapse-prevention trials depend on an assumed event rate for adequate power. Based on the present estimates, eligibility criteria that favor patients with higher Positive and lower Negative scores at randomization, and those that enrich for younger patients, should enrich the sample for relapse events. Because these PANSS Positive and Negative symptom associations operate in opposite directions, combining them strengthens this enrichment: the one-stage model implies an approximately 1.6-fold higher relapse hazard for a patient who is 4 points higher on Positive symptoms and 4 points lower on Negative symptoms than another patient. The opposite profile is more suitable for trials targeting negative symptoms or cognition. These studies require participants to remain clinically stable long enough for change in the primary outcome to be measured; relapse is a competing event that removes participants before assessment is possible. Enrolling patients with prominent negative symptoms and minimal positive symptoms, as the scientific question already requires, may therefore also reduce the attrition that most threatens such trials. This interpretation is consistent with the low rate of symptomatic worsening observed over 12 months in the roluperidone negative symptom program.^11^ Because operational inclusion and exclusion criteria strongly influence who is enrolled,^27^ these results suggest that they may also shape event rates. The modest association between older age and lower relapse risk, equivalent to a 13% reduction per decade, points in the same direction and may also be worth considering when designing such trials.

## Limitations

First, although patients randomized to placebo from five trials were pooled, the resultant sample size was modest, reducing the statistical power for the planned analyses. Second, relapse was defined using a single PANSS-based criterion. Although this criterion was derived against clinician-rated relapse and hospitalization^21^ and allowed harmonization across protocols, it is not identical to the sponsor-or investigator-defined endpoints. Third, the per-point hazard ratios were modest, and the confidence interval for the Positive subscale was wider than that for the Negative subscale and extended slightly below unity. The model-implied ratios for larger score differences are easier to interpret but inherit, and may amplify, this uncertainty. Fourth, although all five trials were double-blind, placebo-controlled antipsychotic withdrawal, relapse-prevention studies that followed stabilization after an acute schizophrenia exacerbation, trial designs and populations were not identical: they differed in baseline severity, double-blind study duration, and visit frequency (Table 1). The main pooled analysis treated them as sharing a common underlying relapse pattern, which may overstate the negative-symptom association. When each trial was analyzed separately and the results were then combined, the estimated negative-symptom effect was about half as large and was no longer statistically significant. With only five trials, each including a limited number of placebo-treated participants, the available data cannot establish which estimate is closer to the truth. Thus, the most cautious conclusion is that the direction of the association is supported, but its magnitude remains uncertain, requiring further validation in future studies.

## Conclusion

Across the placebo arms of five double-blind, placebo-controlled antipsychotic withdrawal relapse-prevention trials, baseline positive and negative symptoms predicted time to schizophrenia relapse in opposite directions and, in the pooled analysis, with nearly equal effect sizes. The protective association for negative symptoms, previously reported in medicated cohorts, was also observed in the pooled analysis of patients receiving no active compound, although it was weaker when trials were analyzed separately, suggesting that it may reflect the course of the illness rather than a secondary effect of treatment. Baseline PANSS Positive and Negative scores may therefore help enrich relapse-prevention trials and reduce relapse-related attrition in negative-symptom and cognition trials, but these uses should be prospectively validated before they are widely incorporated into eligibility criteria.

## Data Availability

All data produced are available online at yoda.yale.edu and vivli.org

## Author Contributions

JR and MD conceived the analysis and designed the study. JR and MSB conducted the statistical analysis. AN and AC contributed to conceptualization and reviewed the results. JR drafted the manuscript. MD, HS, RL and CUC contributed to interpretation of the results and critically revised the manuscript. All authors approved the final version and agree to be accountable for all aspects of the work.

## Acknowledgments

This publication is based on research using data from Eli Lilly and Company that has been made available through Vivli, Inc. Vivli has not contributed to or approved, and is not in any way responsible for, the contents of this publication.

This study, carried out under YODA Project 2025-0120, used data obtained from the Yale University Open Data Access Project, which has an agreement with Johnson & Johnson. The interpretation and reporting of research using this data are solely the responsibility of the authors and do not necessarily represent the official views of the Yale University Open Data Access Project or Johnson & Johnson.

## Data Availability

The individual participant data underlying this analysis are held by Vivli (https://vivli.org) and the YODA Project (https://yoda.yale.edu) and are available to qualified researchers upon application to those platforms. The approved data requests for the present analysis are publicly posted: the Vivli request at https://doi.org/10.25934/PR00011236, and the YODA Project proposal as 2025-0120 at https://yoda.yale.edu 2025-0120 - The YODA Project. Both set out the objective, hypothesis, outcome definitions and analysis plan as submitted before any data were released. Analysis code is available from the corresponding author on reasonable request.

---

**Supplemental Figure 1.**
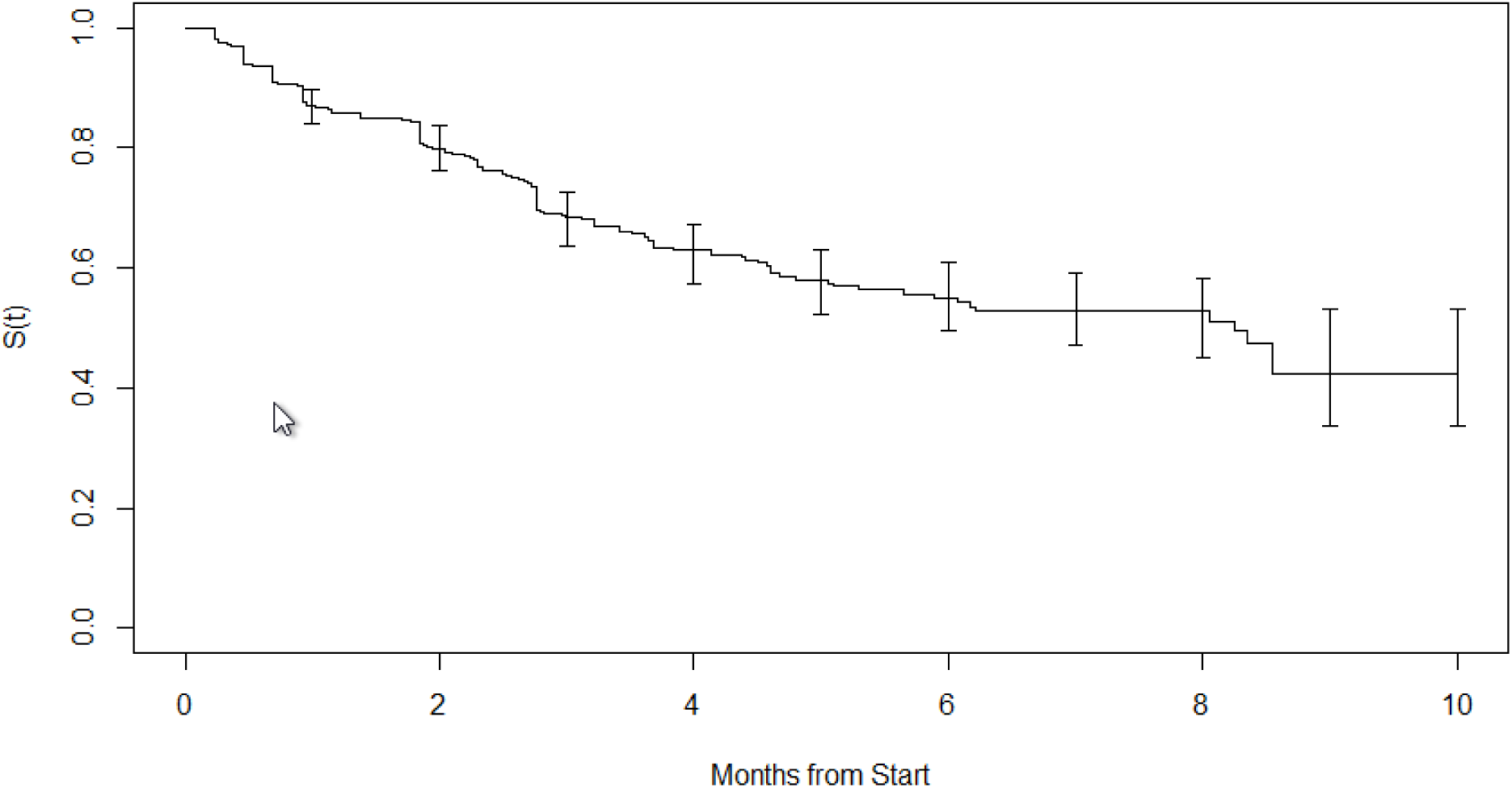
Kaplan-Meier estimate of relapse-free survival for all placebo-randomized participants pooled across the five studies (N = 557). Relapse was defined as an increase of 12 points or more in PANSS total score from double-blind baseline. Vertical bars are 95% confidence intervals at monthly intervals.

**Supplemental Figure 2.**
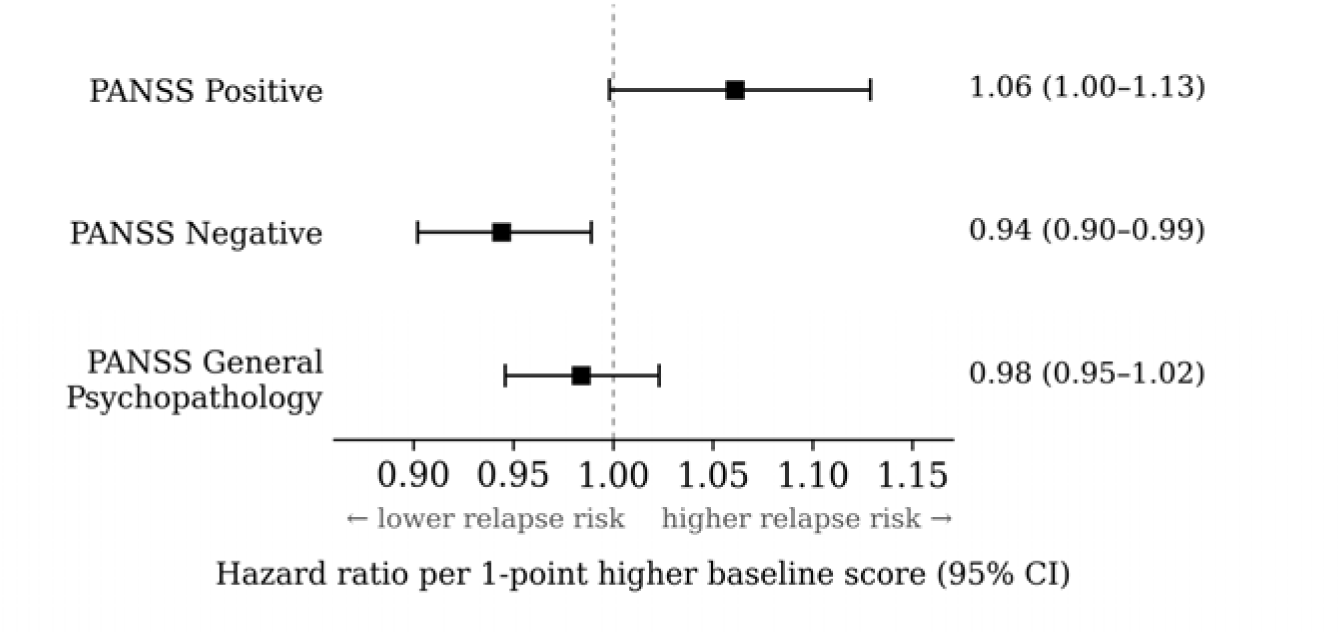
Hazard ratios with 95% confidence intervals for baseline PANSS Positive, Negative and General Psychopathology subscale scores, per 1-point higher score, from the Cox model in Table 2.

## References

1. Ceraso A, Lin JJ, Schneider-Thoma J, et al. Maintenance treatment with antipsychotic drugs for schizophrenia. Cochrane Database Syst Rev. 2020;8(8):CD008016.

2. Davidson M, Carpenter WT Jr. Targeted treatment of schizophrenia symptoms as they manifest, or continuous treatment to reduce the risk of psychosis recurrence. Schizophr Bull. 2024;50(1):14–21.

3. Fountoulakis KN, Moeller HJ, Kasper S, et al. The report of the joint WPA/CINP workgroup on the use and usefulness of antipsychotic medication in the treatment of schizophrenia. CNS Spectr. 2021;26(6):562–586.

4. Solmi M, Murru A, Pacchiarotti I, et al. Safety, tolerability, and risks associated with first-and second-generation antipsychotics: a state-of-the-art clinical review. Ther Clin Risk Manag. 2017;13:757–777.

5. Carbon M, Kane JM, Leucht S, Correll CU. Tardive dyskinesia risk with first-and second-generation antipsychotics in comparative randomized controlled trials: a meta-analysis. World Psychiatry. 2018;17(3):330–340.

6. Correll CU, Schooler NR. Negative symptoms in schizophrenia: a review and clinical guide for recognition, assessment, and treatment. Neuropsychiatr Dis Treat. 2020;16:519–534.

7. Correll CU, Rubio JM, Kane JM. What is the risk-benefit ratio of long-term antipsychotic treatment in people with schizophrenia? World Psychiatry. 2018;17(2):149–160.

8. Rabinowitz J, Levine SZ, Barkai O, Davidov O. Dropout rates in randomized clinical trials of antipsychotics: a meta-analysis comparing first-and second-generation drugs and an examination of the role of trial design features. Schizophr Bull. 2009;35(4):775–788.

9. Schoretsanitis G, Kane JM, Correll CU, Rubio JM. Predictors of lack of relapse after random discontinuation of oral and long-acting injectable antipsychotics in clinically stabilized patients with schizophrenia: a re-analysis of individual participant data. Schizophr Bull. 2022;48(2):296–306.

10. Speyer H, Rabinowitz J, Luthringer R, Tamba BI, Davidson M. Moderate to severe negative symptoms predict low risk of symptoms worsening in schizophrenia patients in CATIE. Schizophr Res. 2026;295:150–155.

11. Rabinowitz J, Staner C, Saoud J, et al. Long-term effects of roluperidone on negative symptoms of schizophrenia. Schizophr Res. 2023;255:9–13.

12. Schneider-Thoma J, Zhu Y, Qin M, et al. Comparative efficacy and tolerability of antidopaminergic and muscarinic antipsychotics for acute schizophrenia: a network meta-analysis of randomised controlled trials indexed in international English and Chinese databases. Lancet. 2026;407(10531):876–891.

13. Rabinowitz J. Predictors of which patients will maintain symptom stability in absence of antipsychotic treatment for schizophrenia. Vivli data request 00011236. 2025. doi:10.25934/PR00011236

14. Rabinowitz J. Identifying predictors of symptom stability in absence of antipsychotic treatment. YODA Project research proposal 2025-0120. 2025. https://yoda.yale.edu

15. Kay SR, Fiszbein A, Opler LA. The Positive and Negative Syndrome Scale (PANSS) for schizophrenia. Schizophr Bull. 1987;13(2):261–276.

16. Kramer M, Simpson G, Maciulis V, et al. Paliperidone extended-release tablets for prevention of symptom recurrence in patients with schizophrenia: a randomized, double-blind, placebo-controlled study. J Clin Psychopharmacol. 2007;27(1):6–14.

17. Hough D, Gopal S, Vijapurkar U, Lim P, Morozova M, Eerdekens M. Paliperidone palmitate maintenance treatment in delaying the time-to-relapse in patients with schizophrenia: a randomized, double-blind, placebo-controlled study. Schizophr Res. 2010;116(2-3):107–117.

18. Janssen Research & Development, LLC. Paliperidone extended release tablets for the prevention of relapse in subjects with schizophrenia: a randomized, double-blind, placebo-controlled, parallel-group study. Clinical study report synopsis. ClinicalTrials.gov identifier NCT01662310; sponsor protocol R076477-SCH-3041 / CR100427. 2013.

19. Berwaerts J, Liu Y, Gopal S, et al. Efficacy and safety of the 3-month formulation of paliperidone palmitate vs placebo for relapse prevention of schizophrenia: a randomized clinical trial. JAMA Psychiatry. 2015;72(8):830–839.

20. Eli Lilly and Company. Olanzapine relapse prevention versus placebo in the treatment of schizophrenia: clinical study report for Study F1D-MC-HGGI. 1999.

21. Siafis S, Brandt L, McCutcheon RA, et al. Relapse in clinically stable adult patients with schizophrenia or schizoaffective disorder: evidence-based criteria derived by equipercentile linking and diagnostic test accuracy meta-analysis. Lancet Psychiatry. 2024;11(1):36–46.

22. Kirkpatrick B, Fenton WS, Carpenter WT, Marder SR. The NIMH-MATRICS consensus statement on negative symptoms. Schizophr Bull. 2006;32(2):214–219.

23. Marder SR, Galderisi S. The current conceptualization of negative symptoms in schizophrenia. World Psychiatry. 2017;16(1):14–24.

24. Nordon C, Rouillon F, Azorin JM, Barry C, Urbach M, Falissard B. Trajectories of antipsychotic response in drug-naive schizophrenia patients: results from the 6-month ESPASS follow-up study. Acta Psychiatr Scand. 2014;129(2):116–125.

25. Austin SF, Mors O, Budtz-Jørgensen E, et al. Long-term trajectories of positive and negative symptoms in first episode psychosis: a 10 year follow-up study in the OPUS cohort. Schizophr Res. 2015;168(1-2):84–91.

26. Wunderink L, van Bebber J, Sytema S, Boonstra N, Meijer RR, Wigman JTW. Negative symptoms predict high relapse rates and both predict less favorable functional outcome in first episode psychosis, independent of treatment strategy. Schizophr Res. 2020;216:192–199.

27. Rabinowitz J, Werbeloff N, Caers I, et al. Negative symptoms in schizophrenia - the remarkable impact of inclusion definitions in clinical trials and their consequences. Schizophr Res. 2013;150(2-3):334–338.

